# All-cause and infection-attributable mortality amongst adults with bloodstream infection – a population-based study

**DOI:** 10.1101/2023.09.29.23296346

**Authors:** Jonathan Underwood, Rowena Griffiths, David Gillespie, Ashley Akbari, Haroon Ahmed

## Abstract

**Background:** Bloodstream infections (BSI), are common, life threatening infections. However, it remains unclear whether deaths following BSI are primarily due to uncontrolled infection or underlying comorbidities. We aimed to determine the overall mortality, infection-attributable mortality, and causes of death for four leading BSI pathogens.

**Methods:** This retrospective cohort study was conducted within the SAIL Databank, containing anonymised population-scale electronic health record data for Wales, UK. We included adults with *Escherichia coli, Klebsiella sp, Pseudomonas aeruginosa* and *Staphylococcus aureus* BSI between 2010-2022 using linked data from Public Health Wales and the Office for National Statistics. 30-day all-cause and sepsis-specific mortality, as a proxy for infection-attributable mortality, were compared using Cox proportional hazards and competing risk regression respectively.

**Findings:** We identified 35,691 adults with BSI. *E. coli* was the most prevalent (59.6%). Adjusted analyses revealed that all organisms had a higher 30-day mortality vs. *E. coli* with MRSA the highest (HR: 2.04 [1.83-2.37], p<0.001).

Cancer was the leading cause of death following BSI for all organisms, particularly deaths occurring between 30-90 days (35.9%). 25.5% of deaths within 30 days involved sepsis. MRSA was associated with the highest sepsis mortality vs. *E. coli* (HR: 2.45 [2.12-2.82], p<0.001). Peak CRP was positively associated with increased sepsis mortality (p<0.001).

**Interpretation:** This population-level study challenges the assumption that most deaths following BSI are directly attributable to uncontrolled infection. Our findings underscore the need for re-evaluating clinical trial design and developing better preventative strategies for BSI.

**Funding:** This work is funded by the Medical Research Council [grant number MR/T023791/1].

## Introduction

Bacteraemias, or bloodstream infections (BSI), are common, life-threatening infections with high mortality rates worldwide.^1^ Whilst the reported 30-day mortality for BSI ranges from 15-30%^1–3^ it is unclear whether these deaths are primarily due to the infection itself or underlying morbidities. The implicit assumption with published BSI mortality figures is that most deaths are directly due to uncontrolled infection and sepsis. However, with modern healthcare and an ageing multimorbid population, many patients with BSI may die of their underlying morbidities, some of which may have precipitated the BSI, rather than of overwhelming infection. For example, in the largest published randomised controlled trial (RCT) of *Staphylococcus aureus* BSI (SAB) only half of deaths were attributed to SAB, with these deaths peaking in the first week.^4^ Further details of the exact causes of death in the remaining 50% of people in this study were not reported, highlighting a wider problem of sparsity of data reporting cause of death after BSI. A Danish population-based study ^5^ examined longer-term (median 1.2 years) causes of death following 25,855 SAB cases from 1992-2014. In total, 22% died of cardiovascular disease, 23% of cancer, and only 8% had a cause of death related to infection. Attributable mortality or sepsis-associated mortality, a reasonable proxy for attributable death from uncontrolled infection, was not reported. A more recent study^6^ from Queensland, Australia, reported mortality and causes of death following 7,061 cases of Gram-negative BSI between 2005-2010. Gastrointestinal malignancy was the commonest cause of death between 0-90 days and 90-365 days and up to four years after infection. This likely reflects many Gram-negative BSI being a consequence of GI malignancy (e.g., cholangitis from an obstructing tumour). Sepsis was reported as the fifth commonest underlying cause of death between 0-90 days accounting for roughly 20% of the total caused by gastrointestinal malignancy. Data were not reported as percentages of total deaths nor stratified by organism, preventing comparison between different organisms.

Detailed patient-level mortality data from Randomised Controlled Trials (RCTs) are conflicting, reflect highly selected populations, and are rarely reported in the main trial publication. Large RCTs of people with drug-resistant Gram-negative BSI, many of whom had advanced cancer, have reported deaths attributable to BSI varying between 0-45%.^7,8^

Understanding causes of death following BSI is essential to determine the infection attributable mortality and to successfully design clinical trials testing therapeutics. Here, we aimed to determine overall and infection attributable mortality and causes of death at different time points following BSI for the four leading BSI pathogens, *Escherichia coli, Klebsiella sp, Pseudomonas aeruginosa (PsA) and Staphylococcus aureus* using high-quality population level linked clinical and microbiological data.

## Methods

### Study design and population

We performed a retrospective cohort study of adults (aged >18 years at the time of BSI) with *Escherichia coli, Klebsiella sp, Pseudomonas aeruginosa (PsA) and Staphylococcus aureus* BSI. These organisms were selected due to their global burden and prevalence. To be eligible participants needed to have a BSI between April 2010-2022 diagnosed via a blood culture collected in a Welsh NHS hospital.

### Data sources

All microbiological samples taken from NHS primary and secondary care services in Wales are processed in United Kingdom Accreditation Services (UKAS) accredited laboratories using standardised methodology. Blood cultures that grew *Escherichia coli, Klebsiella sp, Pseudomonas aeruginosa (PsA) and Staphylococcus aureus* from 1^st^ April 2010 to 31^st^ March 2022 were extracted from Public Health Wales’ data. These data were linked to anonymized individual-level, population-scale, routinely collected electronic health record (EHR) data within the Secure Anonymised Information Linkage (SAIL) Databank.^9^ The linked data-sets included hospital episode data extracted from the Patient Episode Database for Wales (PEDW). C-reactive protein (CRP) measurements were extracted from the Welsh Results Reporting Service (WRRS), which contains data for all laboratory samples processed in NHS laboratories across Wales. Peak CRP, a proxy of the magnitude of the inflammatory response, was determined between the period starting two days before the BSI and ending seven days after, to account for asynchronous blood draws as blood cultures are often taken separately when patients are febrile.

Positive cultures with the same organism within 14 days were considered to be the same BSI. *Staphylococcus aureus* BSI was subdivided by methicillin sensitivity due to national reporting conventions. More than one organism isolated from the same sample was regarded as ‘polymicrobial’ and considered separately. In the case of multiple positive blood cultures, only the most recent was considered for analysis.

Causes of death were determined from linked Office for National Statistics (ONS) records which are based on the death certificate written by the attending doctor. Underlying cause of death was determined using standard ONS methodology based on International Classification of Diseases 10^th^ Revision (ICD-10) and World Health Organisation (WHO) definitions.^10^ We also looked for cases with any mention of sepsis listed as a cause of death as another method of quantifying BSI-attributable deaths. This framework is similar to the ONS method for official reporting of deaths involving COVID-19.^11^ Deaths involving sepsis, hereafter sepsis deaths, were defined using already established ICD-10 code lists (supplementary table m1).^12^ Deaths were grouped according to standard ONS methodology (e.g. malignant neoplasms (cancer) and dementia). Code lists are in supplementary table m2.

### Exposure

The exposure was most recent monomicrobial blood culture that grew *Escherichia coli, Klebsiella sp, Pseudomonas aeruginosa (PsA) or Staphylococcus aureus*. Polymicrobial infections including these organisms were considered separately.

### Outcomes

The primary outcomes were 30-day all-cause mortality and 30 and 90-day cause-specific mortality following BSI. Secondary outcomes included mortality associated with sepsis at 30 days following BSI.

### Covariates

Baseline covariates included age, sex, number of comorbidities, Charlson comorbidity index (CCI) score, Welsh Index of Multiple Deprivation (WIMD) version 2019 as quintiles mapped from Lower-level Super Output Area (LSOA) of residence (administrative authority locality) and electronic frailty index (EFI).^13^ Chosen covariates could all confound the relationship between BSI organism and mortality and were available in a standardised format at the population level.

### Statistical analysis

Demographic characteristics were compared by organism with Kruskal-Wallis rank sum and Pearson’s Chi-squared tests as appropriate. Kaplan-Meier graphs were used to show mortality following bloodstream infection stratified by BSI organism. The log-rank test was used to assess differences in mortality by organism.

In the mortality analyses, associations between BSI organism and 30-day mortality were determined using a Cox proportional hazards model adjusting for age, sex, WIMD, CCI and EFI. Associations between BSI organism and 30-day sepsis death was determined using competing risks regression, a proportional sub-distribution hazards regression model, (with death from a non-sepsis cause as a competing risk) adjusting for age, sex, WIMD, CCI and EFI.

To assess how the magnitude of the inflammatory response affected infection-attributable mortality the association between peak CRP and 30-day sepsis death was determined using competing risks regression (with death from a non-sepsis cause as a competing risk) adjusting for organism, age, sex, WIMD, CCI and EFI. As there are no accepted thresholds for CRP, it was arbitrarily split into five groups: <100; 100-199; 200-299; 300-399; 400+ mg/L.

All statistical analyses were conducted using R v4.1.3 using survival and cmprsk packages.^14^

## Results

35,691 adults with BSI were identified, of whom 21,270 (59.6%) were *E. coli*; 3,488 (9.8%) *Klebsiella sp*.; 6,558 (18.4%) MSSA; 957 (2.7%) MRSA; 1,177 (3.3%) PsA BSI; and 2,241 (6.3%) were polymicrobial. Median age ranged from 69 years for MSSA, to 77 years for MRSA (Table 1). A greater proportion of females were observed in the *E. coli* BSI group (11,345 [53%]), compared with the other BSI groups which contained >60% males. Patients with MRSA BSI tended to have more comorbidities. Peak CRP level was relatively consistent across the different BSIs.

**Table 1.** Baseline demographics.

| Variable | Organism |  |  |  |  |  | p-value<sup>2</sup> |
| --- | --- | --- | --- | --- | --- | --- | --- |
|  | E. coli, N = 21,270<sup>1</sup> | Klebsiella, N = 3,488<sup>1</sup> | MRSA, N = 957<sup>1</sup> | MSSA, N = 6,558<sup>1</sup> | Polymicrobial, N = 2,241<sup>1</sup> | PsA, N = 1,177<sup>1</sup> |  |
| **Age (years)** | 76 (66 – 84) | 74 (64 – 82) | 77 (66 – 85) | 69 (54 – 80) | 74 (63 – 83) | 75 (66 – 83) | <0.001 |
| **Sex** |  |  |  |  |  |  | <0.001 |
| *Female* | 11,345 (53) | 1,331 (38) | 311 (32) | 2,449 (37) | 879 (39) | 451 (38) |  |
| *Male* | 9,925 (47) | 2,157 (62) | 646 (68) | 4,109 (63) | 1,362 (61) | 726 (62) |  |
| **Frailty Rating** |  |  |  |  |  |  | <0.001 |
| *Fit* | 7,346 (35) | 1,201 (34) | 268 (28) | 2,759 (42) | 734 (33) | 409 (35) |  |
| *Mild* | 6,676 (31) | 1,171 (34) | 272 (28) | 2,021 (31) | 773 (34) | 402 (34) |  |
| *Moderate* | 4,993 (23) | 786 (23) | 266 (28) | 1,286 (20) | 529 (24) | 274 (23) |  |
| *Severe* | 2,255 (11) | 330 (9.5) | 151 (16) | 492 (7.5) | 205 (9.1) | 92 (7.8) |  |
| **Charlson Index** | 8 (0 – 18) | 10 (2 – 22) | 14 (5 – 25) | 8 (0 – 19) | 9 (1 – 19) | 11 (4 – 21) | <0.001 |
| **Welsh Index of Multiple Deprivation** |  |  |  |  |  |  | <0.001 |
| 1 | 4,298 (21) | 723 (22) | 193 (21) | 1,569 (25) | 468 (22) | 249 (22) |  |
| 2 | 4,392 (22) | 690 (21) | 193 (21) | 1,373 (22) | 471 (22) | 219 (20) |  |
| 3 | 4,471 (22) | 641 (19) | 195 (22) | 1,247 (20) | 444 (21) | 216 (19) |  |
| 4 | 3,642 (18) | 608 (18) | 171 (19) | 1,019 (16) | 382 (18) | 192 (17) |  |
| 5 | 3,389 (17) | 642 (19) | 146 (16) | 1,002 (16) | 383 (18) | 245 (22) |  |
| *Missing* | 1,078 | 184 | 59 | 348 | 93 | 56 |  |
| **Peak CRP (mg/L)** | 202 (122 – 290) | 199 (119 – 292) | 206 (124 – 295) | 213 (121 – 311) | 200 (119 – 285) | 214 (134 – 314) | <0.001 |
| *Missing* | 1,858 | 367 | 167 | 687 | 209 | 185 |  |

<sup>1</sup>Median (IQR); n (%)
<sup>2</sup>Kruskal-Wallis rank sum test; Pearson's Chi-squared test

Overall 30-day mortality was highest in patients with MRSA BSI (38.7%, 95% CI: 35.7-41.9) and lowest for patients with *E. coli* BSI (18.2%, 17.7-18.8) (Figure 1). 2,523 patients (32.2% of deaths by 30 days) died in two or fewer days after collection of the blood culture that identified BSI – i.e., before identification using traditional culture-based methods. Baseline characteristics by timing of death in the first 30 days are shown in Table 2. Death within two days of BSI was highest for patients with PsA BSI (45.3%, 40.3-50.3) and lowest for patients with MSSA (24.8%, 22.8-26.9). For patients who only ever had one recorded BSI during the study period (n=31,259, 45% E.coli, supplementary table 1) mortality was very similar to the whole cohort (supplementary table 2).

**Table 2.** Characteristics of patients by timing and cause of death.

| Variable | Timing of death |  |  | Cause of death |  |  |
| --- | --- | --- | --- | --- | --- | --- |
|  | Died 0-2 days,<br>N = 2,523<sup>1</sup> | Died 3-30 days,<br>N = 5,314<sup>1</sup> | p-value<sup>2</sup> | Other causes,<br>N = 5,783<sup>1</sup> | Sepsis, N =<br>2,054<sup>1</sup> | p-value<sup>2</sup> |
| **Age (years)** | 78 (68 – 85) | 79 (69 – 86) | <0.001 | 78 (69 – 86) | 79 (70 – 86) | 0.087 |
| **Sex** |  |  | 0.030 |  |  | 0.001 |
| *Female* | 1,126 (45) | 2,234 (42) |  | 2,418 (42) | 942 (46) |  |
| *Male* | 1,397 (55) | 3,080 (58) |  | 3,365 (58) | 1,112 (54) |  |
| **Frailty Rating** |  |  | 0.76 |  |  | 0.15 |
| *Fit* | 784 (31) | 1,641 (31) |  | 1,797 (31) | 628 (31) |  |
| *Mild* | 762 (30) | 1,655 (31) |  | 1,816 (31) | 601 (29) |  |
| *Moderate* | 681 (27) | 1,384 (26) |  | 1,500 (26) | 565 (28) |  |
| *Severe* | 296 (12) | 634 (12) |  | 670 (12) | 260 (13) |  |
| **Charlson Index** | 14 (4 – 24) | 17 (8 – 26) | <0.001 | 16 (8 – 26) | 14 (4 – 23) | <0.001 |
| **Organism** |  |  | <0.001 |  |  | <0.001 |
| *E. coli* | 1,327 (53) | 2,552 (48) |  | 2,888 (50) | 991 (48) |  |
| *Klebsiella* | 250 (9.9) | 568 (11) |  | 649 (11) | 169 (8.2) |  |
| *MRSA* | 105 (4.2) | 266 (5.0) |  | 255 (4.4) | 116 (5.6) |  |
| *MSSA* | 418 (17) | 1,268 (24) |  | 1,234 (21) | 452 (22) |  |
| *Polymicrobial* | 241 (9.6) | 440 (8.3) |  | 479 (8.3) | 202 (9.8) |  |
| *PsA* | 182 (7.2) | 220 (4.1) |  | 278 (4.8) | 124 (6.0) |  |
| **Welsh Index of**<br>**Multiple Deprivation** |  |  | 0.24 |  |  | 0.93 |
| 1 | 509 (22) | 1,048 (21) |  | 1,139 (21) | 418 (21) |  |
| 2 | 507 (22) | 1,121 (22) |  | 1,186 (22) | 442 (23) |  |
| 3 | 508 (22) | 1,056 (21) |  | 1,150 (21) | 414 (21) |  |
| 4 | 410 (18) | 902 (18) |  | 964 (18) | 348 (18) |  |
| 5 | 373 (16) | 913 (18) |  | 956 (18) | 330 (17) |  |
| *Missing* | 216 | 274 |  | 388 | 102 |  |
| **Peak CRP (mg/L)** | 200 (101 – 308) | 236 (149 – 319) | <0.001 | 219 (127 – 310) | 249 (151 – 332) | <0.001 |
| *Missing* | 329 | 501 |  | 625 | 205 |  |

<sup>1</sup>Median (IQR); n (%)
<sup>2</sup>Wilcoxon rank sum test; Pearson's Chi-squared test

**Figure 1.**
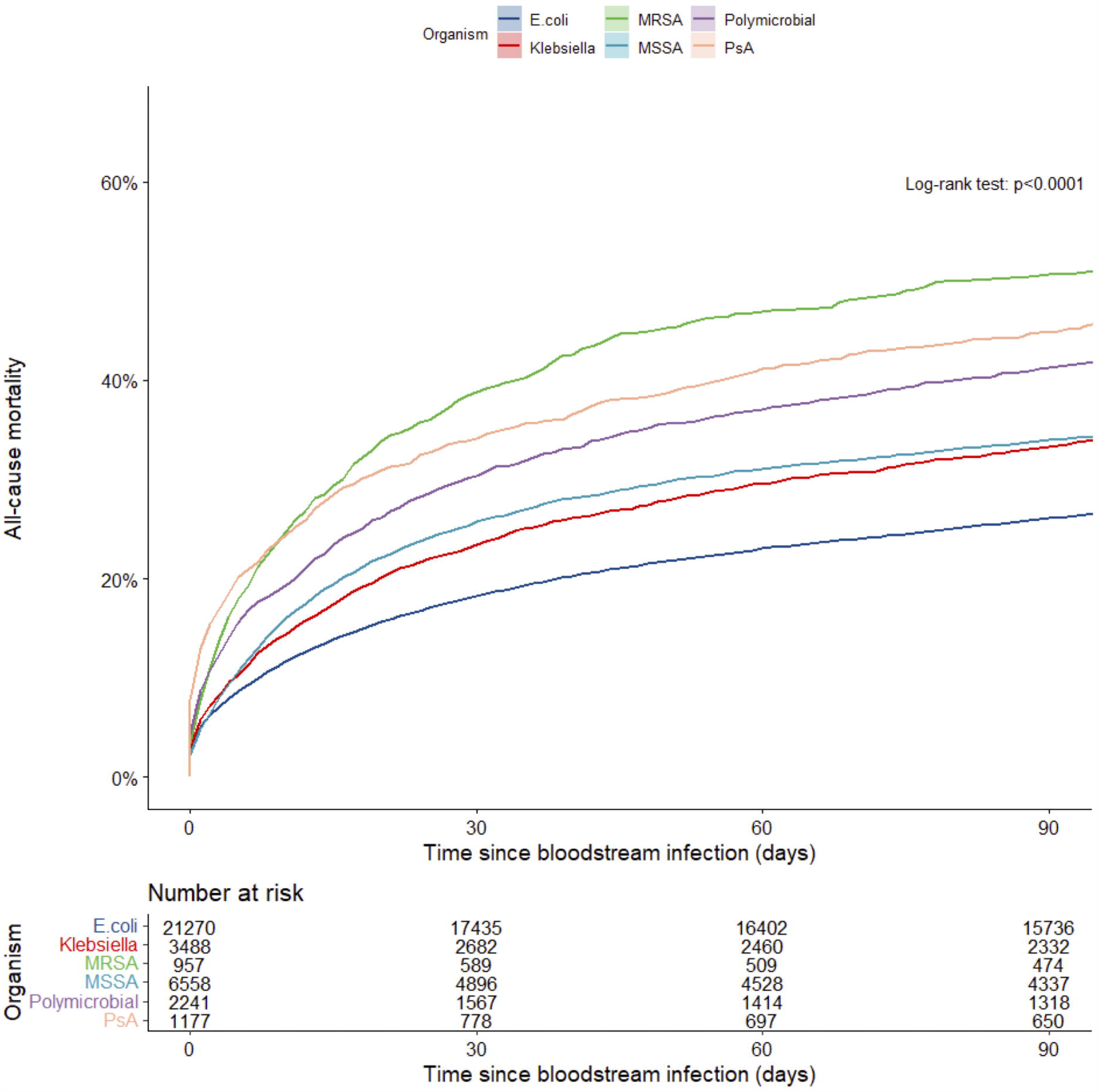
Mortality following bloodstream infection – stratified by organism. Abbreviations: MRSA (methicillin-resistance *Staphylococcus aureus*); MSSA (methicillin-sensitive *Staphylococcus aureus*); PsA (*Pseudomonas aeruginosa*).

After adjusted for age, sex, WIMD, Charlson comorbidity index and frailty, all other organisms were associated with greater 30-day mortality compared with *E. coli* BSI (table 3), with MRSA having the highest mortality (HR [95% CIs]: 2.04 [1.84-2.30]; p<0.001 for all).

**Table 3.** Cox proportional-hazards model for 30-day mortality following BSI stratified by organism adjusted for age, sex, WIMD, Charlson comorbidity index and frailty.

|  | HR<sup>1</sup> | 95% CI<sup>1</sup> | p-value |
| --- | --- | --- | --- |
| **Organism** |  |  |  |
| *E. coli* | — | — |  |
| *Klebsiella* | 1.27 | 1.17 to 1.38 | <0.001 |
| *MRSA* | 2.02 | 1.78 to 2.28 | <0.001 |
| *MSSA* | 1.61 | 1.51 to 1.71 | <0.001 |
| *Polymicrobial* | 1.88 | 1.72 to 2.05 | <0.001 |
| *PsA* | 1.91 | 1.71 to 2.15 | <0.001 |

<sup>1</sup>HR = Hazard Ratio, CI = Confidence Interval

### Causes of death

For all organisms, cancer was the leading underlying cause of death following BSI (25.2% of deaths by 30 days and 35.9% occurring between 30-90 days, Figure 2). For *E. coli* BSI, cancer was the underlying cause of death for 26.9% and 38.8% of deaths occurring up to 30-days and between 30-90 days respectively. The leading specific underlying causes of deaths by 30-days were mainly due to infective causes with urinary tract infection the most common diagnosis. However, after 30-days many deaths were caused by dementia and other non-communicable diseases. For *Klebsiella sp*. BSI, pancreatic cancer was the leading specific cause of death between 30-90 days with malignancy the underlying cause of death in 29.8% and 42.7% occurring up to 30-days and 30-90 days respectively. There were fewer cases of PsA BSI and therefore fewer deaths. However, underlying causes of death followed a similar pattern whereby deaths occurring up to 30-days after BSI were predominantly of infective causes whereas those occurring 30-90 days after BSI were dominated by non-communicable diseases - particularly cancer (31.3% by 30-days and 35.0% 30-90 days). For MSSA BSI, ischaemic heart disease was the second commonest specific underlying cause of death by 30-days and most common between 30-90 days. Cancer was the underlying cause of death in 17.6% by 30-days and 23.6% between 30-90 days. For MRSA, sepsis was the leading specific underlying cause of death at both 30 and 30-90 days. Malignancy was responsible for relatively fewer deaths (16.2% up to 30 days, 20.2% between 30-90 days). Overall, dementia and cancer were leading causes of death between 30-90 days accounting for 6.9% and 35.9% respectively. Cancer was also the leading underlying cause of very early (within two days) deaths after BSI (supplementary figure 1.)

**Figure 2.**
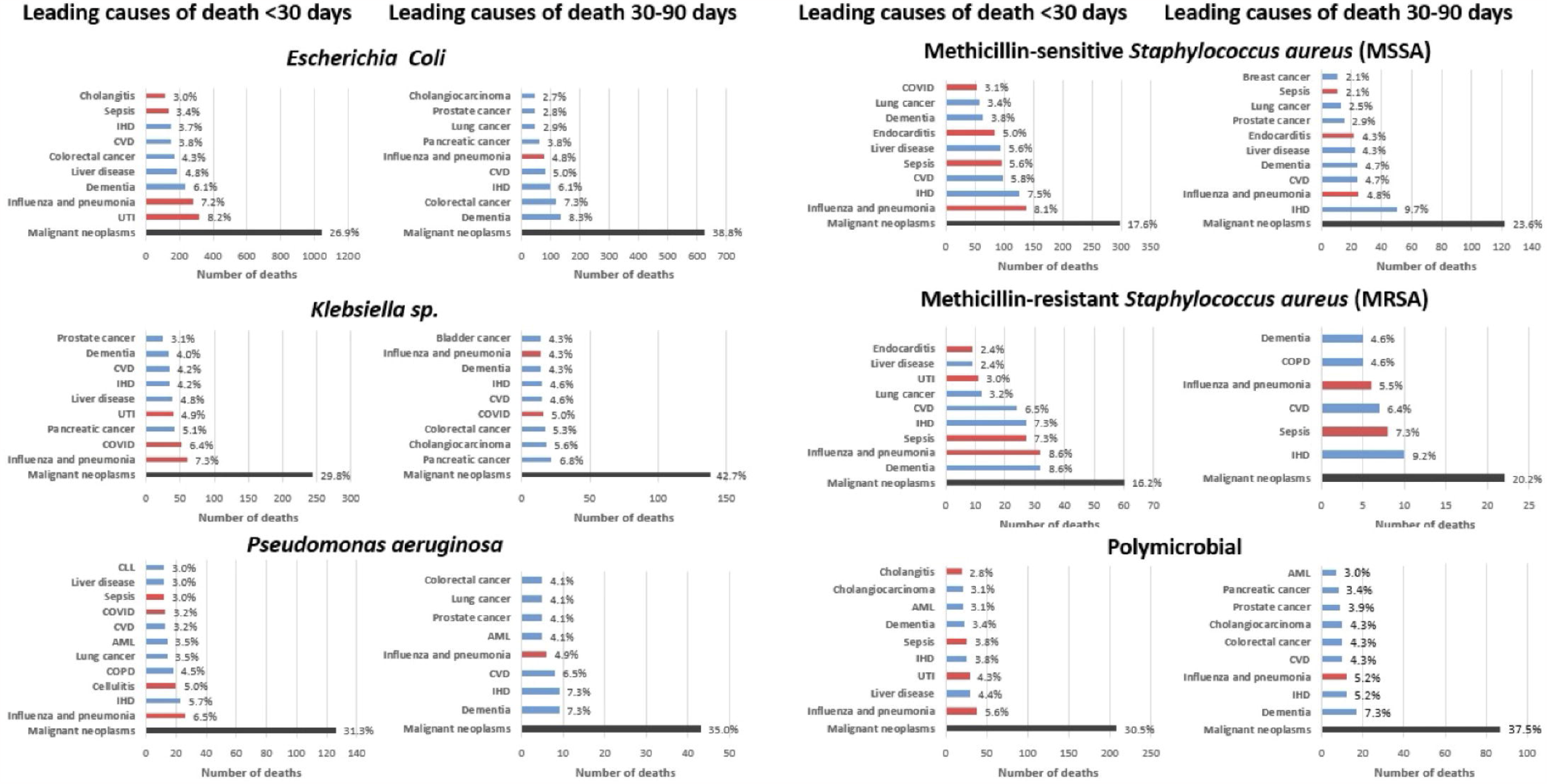
Underlying cause of death by organism. Red bars indicate infective causes. Causes with fewer than five deaths not displayed for SAIL information governance reasons.

### Deaths involving sepsis

Of those who died within 30 days of BSI, 1,505 (18.7%) had sepsis listed as the primary cause of death and 2,054 (25.5%) had sepsis listed as a primary or secondary cause of death. The proportion of deaths due to sepsis reduced over time (figure 3). Median survival was shorter for sepsis versus non-sepsis deaths (7 [7-8] days vs 45 [43-47] days) and a greater proportion of sepsis deaths occurred within two days of BSI (38% vs 29%, p<0.001). Deaths from sepsis as a proportion of deaths by 30-days following BSI varied by organism and were highest for MRSA (31.3% [26.7-36.3]%) and lowest for *Klebsiella sp* BSI (20.7% [17.9-23.6]%). Using a competing risk analysis, 30-day sepsis mortality was significantly greater for MSSA, MRSA, PsA and polymicrobial versus *E. coli* BSI (all p<0.001, Table 4) with MRSA again associated with the highest mortality (HR: 2.55 [2.10-3.10], p<0.001). In an exploratory analysis, the magnitude of the inflammatory response following BSI, measured by peak CRP, was associated with 30-day sepsis mortality. Compared to those with a peak CRP of <100 mg/L having a peak CRP >400mg/L was associated with a higher mortality (HR: 3.00 [2.49-3.62], Table 5).

**Table 4.**
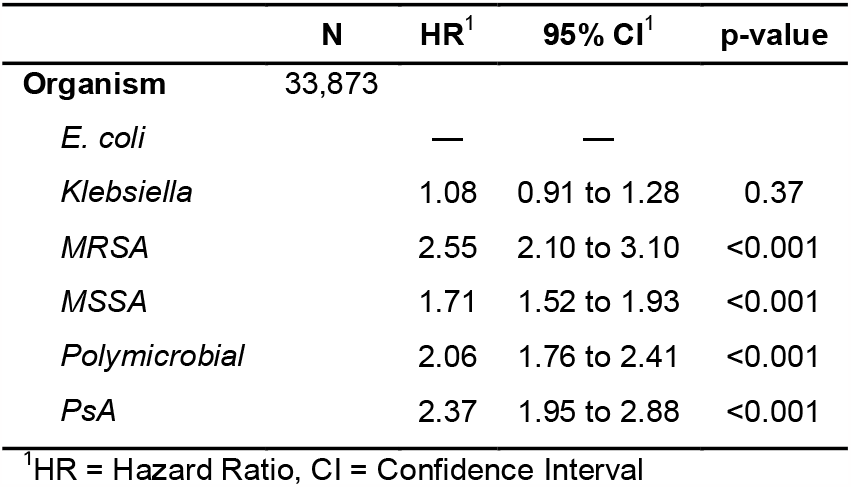
Competing risk regression for 30-day sepsis mortality following BSI stratified by organism adjusted for age, sex, WIMD, Charlson comorbidity index and frailty.

|  | N | HR<sup>1</sup> | 95% CI<sup>1</sup> | p-value |
| --- | --- | --- | --- | --- |
| **Organism** | 33,873 |  |  |  |
| *E. coli* |  | — | — |  |
| *Klebsiella* |  | 1.08 | 0.91 to 1.28 | 0.37 |
| *MRSA* |  | 2.55 | 2.10 to 3.10 | <0.001 |
| *MSSA* |  | 1.71 | 1.52 to 1.93 | <0.001 |
| *Polymicrobial* |  | 2.06 | 1.76 to 2.41 | <0.001 |
| *PsA* |  | 2.37 | 1.95 to 2.88 | <0.001 |

<sup>1</sup>HR = Hazard Ratio, CI = Confidence Interval

**Table 5.** Competing risk regression for 30-day sepsis mortality following BSI stratified by peak CRP adjusted for organism, age, sex, WIMD, Charlson comorbidity index and frailty.

|  | N | HR (95% CI)<sup>1</sup> | p-value |
| --- | --- | --- | --- |
| **Peak CRP (mg/L)** | 30,553 |  |  |
| *Under 100* |  | — |  |
| *100-200* |  | 1.10 (0.94 to 1.29) | 0.24 |
| *200-300* |  | 1.49 (1.27 to 1.74) | <0.001 |
| *300-400* |  | 2.22 (1.89 to 2.61) | <0.001 |
| *400+* |  | 3.00 (2.49 to 3.62) | <0.001 |

<sup>1</sup>HR = Hazard Ratio, CI = Confidence Interval

**Figure 3.**
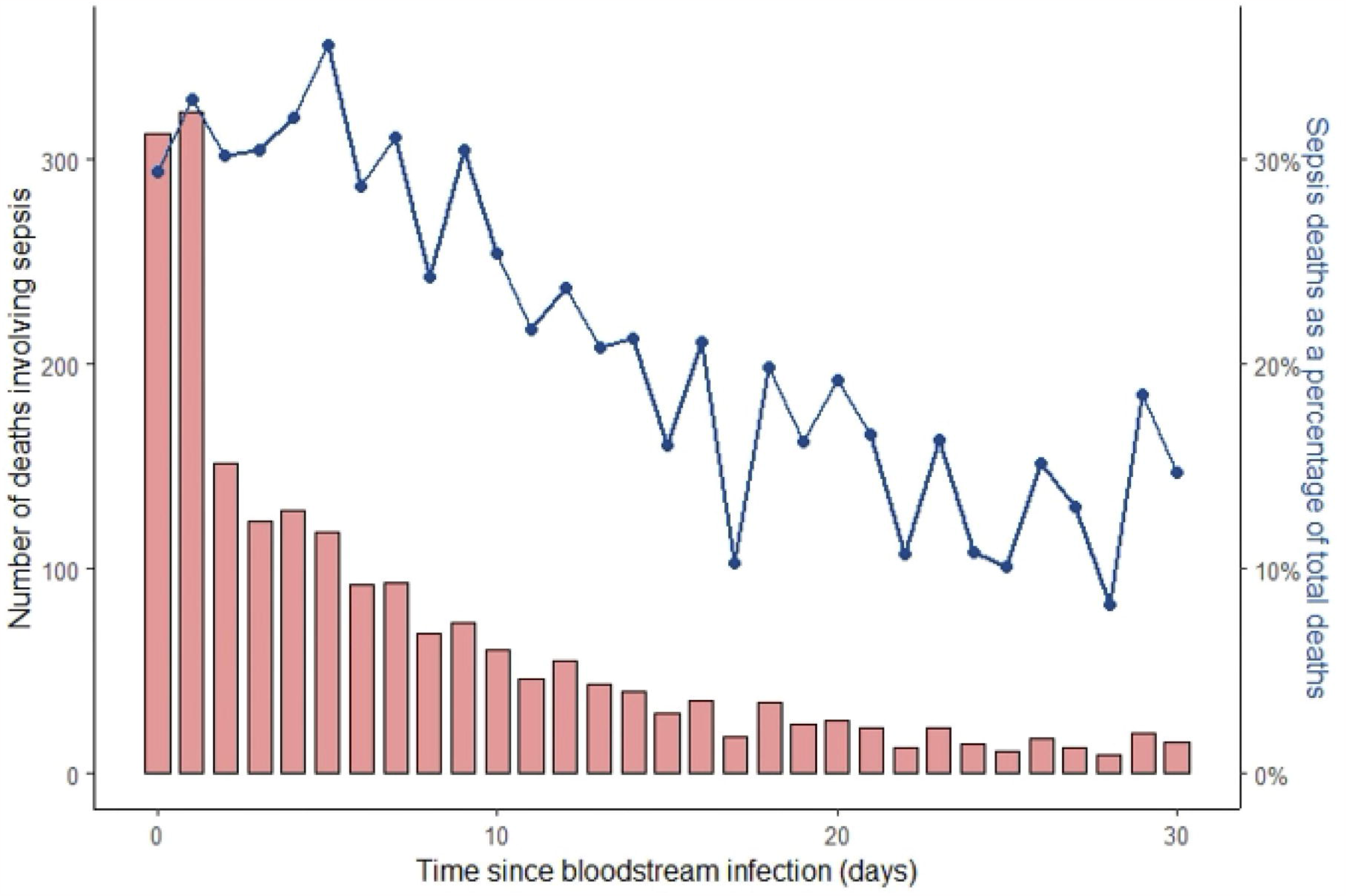
Timing of deaths involving sepsis within 30 days of bloodstream infection.

### Sensitivity analyses

For patients with *E. coli* BSI urinary tract infection was the most common underlying infective cause of death within 30 days. Sepsis was reportedly involved in 48.3% of these cases. There were no differences in potentially confounding factors, peak CRP or days survived between those involving versus those not involving sepsis (supplementary table s3). Similarly for patients with MSSA BSI and endocarditis as the underlying cause of death within 30 days 23.8% reportedly involved sepsis. Again, there were no differences in potentially confounding factors, peak CRP or days survived between those involving versus those not involving sepsis (supplementary table s4).

## Discussion

This national cohort study provides a comprehensive description of mortality, timing, and causes of death following common BSI. We found that many deaths post-BSI are not due to uncontrolled infection but are caused by underlying conditions like cancer, especially in the sub-acute phase - beyond 30 days after BSI.

To our knowledge this is the largest study detailing causes of death following BSI. These data support earlier observational work suggesting that non-infective causes, and in particular cancer, are responsible for most deaths following BSI, particularly for Gram negative pathogens.^5,6^ Pancreatic cancer and cholangiocarcinoma were common underlying causes of death in patients with *E. coli, Klebsiella* and polymicrobial BSI. This is unsurprising given these cancers can cause biliary obstruction leading to biliary stasis, cholangitis and BSI with enteric organisms. The BSI is amenable to curative treatment with antibiotics and supportive care, but the underlying malignancy is not.

Our finding that about a quarter of deaths are directly attributable to BSI is a little higher than previous, smaller observational studies where infection was the reported cause of death in 8-20%.^5,6^ Randomised controlled trials (RCTs) may provide more detailed information on a smaller, selected population, but selection bias may limit their generalisation. In the largest published RCT of SAB, ARREST, half of all deaths were attributed to SAB.^4^ This contrasts sharply with the largest published RCT of ceftriaxone resistant *E. coli* and *Klebsiella* pneumoniae BSI, MERINO,^15^ where no participants died of their index infection. We believe our findings, based on comprehensive, unselected population level data, are robust and provide a reliable, contemporaneous estimate of the infection-attributable mortality of BSI for these common organisms in high-income settings where the prevalence of antimicrobial resistance is relatively low.

We found differential mortality by organism, with MRSA and PsA BSI associated with higher mortality, replicating earlier work.^16–19^ Given differences in underlying causes of death, these differences are likely multifactorial and due to patient factors such as underlying disease (e.g. acute myeloid leukaemia being more common in patients with PsA BSI) and not just intrinsic pathogenicity and antimicrobial resistance. Nonetheless, our competing risks analysis with deaths involving sepsis as the primary outcome suggests that infection-attributable mortality is genuinely higher with these pathogens urging the need for more trials of novel and combination antibiotics in patients with these pathogens.

We found the magnitude of the inflammatory response, quantified with peak CRP within seven days of BSI, was associated with sepsis mortality. Elevated CRP has been associated with overall mortality in BSI previously, with Gram-negative infections reportedly having higher initial concentrations.^16,20,21^ Our robust analyses confirm these earlier findings. Is the magnitude of the inflammatory response causally related to sepsis death? These data cannot answer that question, especially given we do not have treatment data, but it is plausible that infection-triggered inflammation has a destabilising effect on underlying morbidities. However, given the magnitude of the hazard ratio comparing patients with higher versus lower peak CRP, understanding immunopathogenesis is essential to further reduce BSI mortality. Current BSI trials almost exclusively focus on pathogen-directed therapy. Immunomodulation has been shown to be successful in other life-threatening infectious diseases, most notably COVID.^22,23^ Addressing the host-response to BSI in addition to pathogen-directed therapy may reduce mortality and morbidity and should be explored further.

### Strengths and weaknesses

The main strengths of our study are the size of the study population, completeness of data in SAIL and historical microbiological data, allowing an unselected cohort of all adults in Wales with BSI. The main limitation of this study is reliability of death certification for accurately identifying cause of death. To mitigate this, we determined deaths involving sepsis as a proxy for deaths directly attributable to BSI. This also relies on accurate death certification. However, usual practice in the UK is for significant microbiological results, such as BSI, to be telephoned promptly to the attending clinical team. As such, BSI would have been identified in almost every patient by the time of death certification. Therefore, it is reasonable to assume that the treating clinician would put this, sepsis, or a specific infection syndrome, such as endocarditis, as the cause of death if it had contributed to death. This contrasts with other potential infection syndromes, such as pneumonia, where the pathogen is often unidentified and there is more scope for misdiagnosis (e.g., cardiac failure or pulmonary embolus) particularly if death occurs shortly after presentation. Our sensitivity analyses show the limitations of using deaths involving sepsis as a proxy for BSI attributable mortality. Approximately half of patients with an underlying cause of death of urinary tract infection did not have a sepsis code recorded on the death certificate. This observation is hard to reconcile without accepting that death certification ascertainment of sepsis is incomplete (i.e., urinary tract infection is the sole listed cause of death without further detail) and therefore deaths involving sepsis recorded here should be considered a reasonable lower bound of the true number of deaths attributable to uncontrolled infection. A further complication is the changing definition of sepsis over the study period as Sepsis-3 definitions were published in 2016.^24^

Other significant limitations include lack of data regarding the infection syndrome associated with BSI, physiological and other data to confirm the presence of sepsis. Other limitations include lack of detailed antimicrobial resistance data, treatment, residual confounding in adjusted analyses and that causal inference with respect to mortality can only be inferred from an observational study with caution. However, given the large effect sizes observed after adjusting for potential confounders, any bias would have to be large to completely account for our findings. Additionally, missing data was limited to 5% or fewer of the cohort and is unlikely to influence associations.

### Implications for future study

These data have potentially significant implications for clinical trials of BSI. Our data suggest that pathogen directed clinical trials relying on organism identification prior to enrolment will not recruit a significant proportion of people who die of BSI as at least a third are dead or moribund by the time of organism identification. Therefore, these trials will not be representative of all patients with BSI and may be underpowered to detect mortality differences. In the largest published RCT of SAB (6% of whom had MRSA), ARREST, mortality at 12-weeks was 15%.^26^ Half of deaths were attributed to SAB with the majority occurring within the first two weeks.^4^ Using our national sample (14% of whom had MRSA), 30-day overall mortality was nearly double this at 28%. Underestimated mortality post-SAB is not limited to randomised controlled trials. In pooled observational data of SAB (21% MRSA), 90-day mortality was 29% compared with 38% here.^19^ Similarly, in a large trial of drug resistant *E. coli* and *Klebsiella pneumoniae* BSI, MERINO, where mortality may be expected to be higher than usual, overall 30-day mortality was 8%.^15^ This is much lower than the overall 30-day mortality we observed of 20%, again suggesting selection bias. Of the 30 patients who died in this study, 14 (47%) died of malignancy with the remainder dying of other morbidities, particularly liver disease, <u>with none dying of the index BSI</u>.^27^ These data suggest that many people who die of their BSI are not enrolled in clinical trials.

What should we do given these results? Overall mortality as the primary outcome in clinical trials of BSI has the advantage of being unbiased compared with BSI-attributed mortality, although this can be mitigated with blinding and independent panels of experts to attribute cause of death. Analogous to progression-free survival in cancer trials in BSI a composite endpoint of clinical/microbiological recurrence or death, similar to that used in ARREST, may be advantageous. However, given BSI attributable death rates decay significantly over the first two weeks following BSI we advocate 30-rather than 90-day composite endpoints are used.

Earlier identification of culprit pathogens in patients presenting with infection syndromes, using novel technologies, will be key to facilitate earlier enrolment in pathogen specific clinical trials. Alternatives to this approach are novel trial designs that enrol patients with defined life-threatening infection syndromes (e.g. suspected sepsis or urinary tract infection with sepsis) at initial presentation and initially test different empirical therapies with subsequent stratification by organism/resistance mechanism. Key to this would be using a different model of consent such as deferred consent to allow early, less-biased recruitment of patients with life-threatening infections. This model been used successfully used in clinical trials of out-of-hospital cardiac arrest and in patients presenting with suspected sepsis.^28,29^ Given early sepsis mortality and comorbid disease accounting for most later deaths preventative strategies including vaccination are likely to be important for reducing the overall disease burden associated with BSI.

## Conclusion

BSI are common and herald death in a significant proportion. However, sepsis is the direct cause of death in only a quarter of patients, usually occurring in the first few days of infection. Preventative approaches, rapid diagnostics and novel trial design are needed to significantly impact mortality in the future.

## Supporting information

Supplementary methods and results

## Data Availability

This project uses anonymised individual-level data sources held within the Trusted Research Environment provided by the SAIL Databank at Swansea University, Swansea, UK. All proposals to use SAIL data are subject to review by the independent Information Governance Review Panel (IGRP). As such, data is not available for sharing without explicit permission by the IGRP.

## Ethics

This project uses anonymised individual-level data sources held within the Trusted Research Environment provided by the SAIL Databank at Swansea University, Swansea, UK. All proposals to use SAIL data are subject to review by the independent Information Governance Review Panel (IGRP). This work was approved under proposal number 0923 after careful considerations by IGRP Panel.

