## Supplementary methods and results for "All-cause and infection-attributable mortality amongst adults with bloodstream infection – a population-based study"

### Supplementary data

#### Methods

Table m1. ICD-10 codes used to identify sepsis as cause of death.

*ICD 10 codes containing sepsis/septicaemia or SIRS*

| ALT_CODE | DESCRIPTION |
|---|---|
| A021 | Salmonella sepsis |
| A207 | Septicaemic plague |
| A227 | Anthrax sepsis |
| A267 | Erysipelothrix sepsis |
| A327 | Listerial sepsis |
| A40 | Streptococcal sepsis |
| A400 | Sepsis due to streptococcus, group A |
| A401 | Sepsis due to streptococcus, group B |
| A402 | Sepsis due to streptococcus, group D |
| A403 | Sepsis due to Streptococcus pneumoniae |
| A408 | Other streptococcal sepsis |
| A409 | Streptococcal sepsis, unspecified |
| A41 | Other sepsis |
| A410 | Sepsis due to Staphylococcus aureus |
| A411 | Sepsis due to other specified staphylococcus |
| A412 | Sepsis due to unspecified staphylococcus |
| A413 | Sepsis due to Haemophilus influenzae |
| A414 | Sepsis due to anaerobes |
| A415 | Sepsis due to other Gram-negative organisms |
| A418 | Other specified sepsis |
| A419 | Sepsis, unspecified |
| A427 | Actinomycotic sepsis |
| B377 | Candidal sepsis |
| O85X | Puerperal sepsis |
| R65 | Systemic Inflammatory Response Syndrome [SIRS] |
| R650 | Systemic Inflammatory Response Syndrome of infectious origin without organ failure |
| R651 | Systemic Inflammatory Response Syndrome of infectious origin with organ failure |
| R652 | Systemic Inflammatory Response Syndrome of non-infectious origin without organ failure |
| R653 | Systemic Inflammatory Response Syndrome of non-infectious origin with organ failure |
| R659 | Systemic Inflammatory Response Syndrome, unspecified |

Table m2. Leading causes of death – ONS groupings of ICD-10 codes.

| Table 1: Leading causes of death in England and Wales (revised 2016) | |
|---|---|
| ICD-10 codes | Cause of death groups |
| A00–A09 | Intestinal infectious diseases |
| A15–A19, B90 | Tuberculosis |
| A20, A44, A75–A79, A82–A84, A85.2, A90–A98, B50–B57 | Vector–borne diseases and rabies |
| A33–A37, A49.2, A80, B01, B02, B05, B06, B15, B16, B17.0, B18.0, B18.1, B26, B91, G14 | Vaccine-preventable diseases[1] |
| A39, A87, G00–G03 | Meningitis and meningococcal infection |
| A40–A41 | Septicaemia |
| B20–B24 | Human immunodeficiency virus [HIV] disease |
| C00–C97 | Malignant neoplasms |
| C15 | Malignant neoplasm of oesophagus |
| C16 | Malignant neoplasm of stomach |
| C18–C21 | Malignant neoplasm of colon, sigmoid, rectum and anus |
| C22 | Malignant neoplasm of liver and intrahepatic bile ducts |
| C23-C24 | Malignant neoplasm of gallbladder and other parts of biliary tract |
| C25 | Malignant neoplasm of pancreas |
| C32 | Malignant neoplasm of larynx |
| C33-C34 | Malignant neoplasm of trachea, bronchus and lung |
| C40–C41 | Malignant neoplasms of bone and articular cartilage |
| C43-C44 | Melanoma and other malignant neoplasms of skin |
| C50 | Malignant neoplasm of breast |
| C53–C55 | Malignant neoplasm of uterus |
| C56 | Malignant neoplasm of ovary |
| C61 | Malignant neoplasm of prostate |
| C64 | Malignant neoplasm of kidney, except renal pelvis |
| C67 | Malignant neoplasm of bladder |
| C71 | Malignant neoplasm of brain |
| C81–C96 | Malignant neoplasms, stated or presumed to be primary of lymphoid, haematopoietic and related tissue |
| D00–D48 | In situ and benign neoplasms, and neoplasms of uncertain or unknown behaviour |
| E10–E14 | Diabetes |
| D50–D53, E40–E64 | Malnutrition, nutritional anaemias and other nutritional deficiencies |
| E86–E87 | Disorders of fluid, electrolyte and acid–base balance (incl. dehydration) |
| F01, F03, G30 | Dementia and Alzheimer disease |
| F10–F19 | Mental and behavioural disorders due to psychoactive substance use |
| G10–G12 | Systemic atrophies primarily affecting the central nervous system |
| G20 | Parkinson disease |
| G40-G41 | Epilepsy and status epilepticus |
| G80–G83 | Cerebral palsy and other paralytic syndromes |

| | |
|---|---|
| I05–I09 | Chronic rheumatic heart diseases |
| I10–I15 | Hypertensive diseases |
| I20–I25 | Ischaemic heart diseases |
| I26–I28 | Pulmonary heart disease and diseases of pulmonary circulation |
| I34–I38 | Nonrheumatic valve disorders and endocarditis |
| I42 | Cardiomyopathy |
| I46 | Cardiac arrest |
| I47–I49 | Cardiac arrhythmias |
| I50–I51 | Heart failure and complications and ill–defined heart disease |
| I60–I69 | Cerebrovascular diseases |
| I70 | Atherosclerosis |
| I71 | Aortic aneurysm and dissection |
| J00–J06, J20–J22 | Acute respiratory infections other than influenza and pneumonia |
| J09–J18 | Influenza and pneumonia |
| J40–J47 | Chronic lower respiratory diseases |
| J80–J84 | Pulmonary oedema and other interstitial pulmonary diseases |
| J96 | Respiratory failure |
| K35–K46, K56 | Appendicitis, hernia and intestinal obstruction |
| K70–K76 | Cirrhosis and other diseases of liver |
| M00–M99 | Diseases of the musculoskeletal system and connective tissue |
| N00–N39 | Diseases of the urinary system |
| O00–O99 | Pregnancy, childbirth and the puerperium |
| P00–P96 | Certain conditions originating in the perinatal period |
| Q00–Q99 | Congenital malformations, deformations and chromosomal abnormalities |
| V01–X59 | Accidents |
| V01–V89 | Land transport accidents |
| W00–W19 | Accidental falls |
| W32–W34 | Non-intentional firearm discharge |
| W65–W74 | Accidental drowning and submersion |
| W75–W84 | Accidental threats to breathing |
| X40–X49 | Accidental poisoning |
| X60–X84, Y10–Y34 | Suicide and injury/poisoning of undetermined intent[2] |
| U50.9, X85–Y09, Y87.1 | Homicide and probable homicide |
| R00-R99 | Symptoms, signs and ill-defined conditions |
| Source: Office for National Statistics | |
| Notes: | |
| 1. Excluding meningitis and meningococcal diseases (A39), sepsis due to haemophilus influenzae (A41.3), rabies (A82), certain mosquito-borne diseases (A83) and yellow fever (A95). | |
| 2. In England and Wales, a conclusion of suicide cannot be returned for children under the age of 10 years. | |

### Results

#### Table s1. BSI demographics by number of BSI episodes

| | Number of BSI | | |
|---|---|---|---|
| **Variable** | **Single infection**, N = 31,259[1] | **Multiple infections**, N = 4,432[1] | **p-value**[2] |
| **Age (years)** | 75 (63 – 83) | 75 (64 – 83) | 0.32 |
| **Sex** | | | <0.001 |
| *Female* | 14,911 (48) | 1,855 (42) | |
| *Male* | 16,348 (52) | 2,577 (58) | |
| **Frailty Rating** | | | <0.001 |
| *Fit* | 11,322 (36) | 1,395 (31) | |
| *Mild* | 9,867 (32) | 1,448 (33) | |
| *Moderate* | 7,009 (22) | 1,125 (25) | |
| *Severe* | 3,061 (9.8) | 464 (10) | |
| **Charlson Index** | 8 (0 – 19) | 9 (2 – 19) | <0.001 |
| **Organsim** | | | <0.001 |
| *E. coli* | 19,259 (62) | 2,011 (45) | |
| *Klebsiella* | 3,312 (11) | 176 (4.0) | |
| *MRSA* | 904 (2.9) | 53 (1.2) | |
| *MSSA* | 6,070 (19) | 488 (11) | |
| *Polymicrobial* | 597 (1.9) | 1,644 (37) | |
| *PsA* | 1,117 (3.6) | 60 (1.4) | |
| **Welsh Index of Multiple Deprivation** | | | 0.45 |
| *1* | 6,521 (22) | 979 (23) | |
| *2* | 6,411 (22) | 927 (22) | |
| *3* | 6,337 (21) | 877 (21) | |
| *4* | 5,269 (18) | 745 (18) | |
| *5* | 5,101 (17) | 706 (17) | |
| *Missing* | 1,620 | 198 | |
| **Peak CRP (mg/L)** | 205 (122 – 296) | 197 (119 – 281) | <0.001 |
| *Missing* | 3,119 | 354 | |

[1]Median (IQR); n (%)
[2]Wilcoxon rank sum test; Pearson's Chi-squared test

#### Table s2. BSI mortality by organism for patients with only one recorded BSI.

| | Status at 30-days | | |
|---|---|---|---|
| | Alive | Dead | **Total** |
| **Organism** | | | |
| *E. coli* | 15,810 (82%) | 3,449 (18%) | 19,259 (100%) |
| *Klebsiella* | 2,540 (77%) | 772 (23%) | 3,312 (100%) |
| *MRSA* | 549 (61%) | 355 (39%) | 904 (100%) |
| *MSSA* | 4,484 (74%) | 1,586 (26%) | 6,070 (100%) |
| *Polymicrobial* | 403 (68%) | 194 (32%) | 597 (100%) |
| *PsA* | 739 (66%) | 378 (34%) | 1,117 (100%) |
| **Total** | 24,525 (78%) | 6,734 (22%) | 31,259 (100%) |

Table s3. E. coli BSI deaths with UTI as underlying cause of infection stratified by whether sepsis listed as a cause of death anywhere on the death certificate

| Characteristic | **Sepsis not mentioned**, N = 163[1] | **Sepsis**, N = 155[1] | p-value[2] |
|---|---|---|---|
| **Age** | 84 (78 – 90) | 84 (78 – 90) | >0.99 |
| **Days survived** | 4.0 (2.0 – 11.0) | 5.0 (1.5 – 10.0) | 0.96 |
| **Sex** | | | 0.38 |
| Female | 75 (46) | 79 (51) | |
| Male | 88 (54) | 76 (49) | |
| **Frailty category** | | | 0.86 |
| Fit | 31 (19) | 26 (17) | |
| Mild | 40 (25) | 44 (28) | |
| Moderate | 57 (35) | 54 (35) | |
| Severe | 35 (21) | 31 (20) | |
| **CHARLSON_INDEX** | 14 (3 – 22) | 14 (4 – 24) | 0.65 |
| **WIMD** | | | 0.21 |
| 1 | 31 (21) | 47 (32) | |
| 2 | 29 (19) | 23 (16) | |
| 3 | 38 (26) | 35 (24) | |
| 4 | 24 (16) | 24 (16) | |
| 5 | 27 (18) | 18 (12) | |
| Unknown | 14 | 8 | |
| **Peak CRP (mg/L)** | 252 (160 – 336) | 262 (145 – 320) | 0.41 |
| Unknown | 12 | 20 | |

[1]Median (IQR); n (%)
[2]Wilcoxon rank sum test; Pearson's Chi-squared test

Table s4. MSSA BSI deaths with endocarditis as the underlying cause of infection stratified by whether sepsis listed as a cause of death anywhere on the death certificate

| Characteristic | **Sepsis not mentioned**, N = 64[1] | **Sepsis**, N = 20[1] | p-value[2] |
|---|---|---|---|
| **Age (years)** | 77 (70 – 83) | 74 (67 – 84) | 0.75 |
| **Days survived** | 13 (9 – 22) | 11 (6 – 22) | 0.53 |
| **Sex** | | | 0.71 |
| Female | 35 (55) | 10 (50) | |
| Male | 29 (45) | 10 (50) | |
| **Frailty category** | | | 0.89 |
| **CHARLSON_INDEX** | 13 (2 – 23) | 18 (8 – 22) | 0.57 |
| **WIMD** | | | 0.84 |
| **Peak CRP (mg/L)** | 264 (189 – 334) | 288 (166 – 332) | 0.87 |
| Unknown | <5 | <5 | |

[1]Median (IQR); n (%)
[2]Wilcoxon rank sum test; Pearson's Chi-squared test; Fisher's exact test

Figure s1.

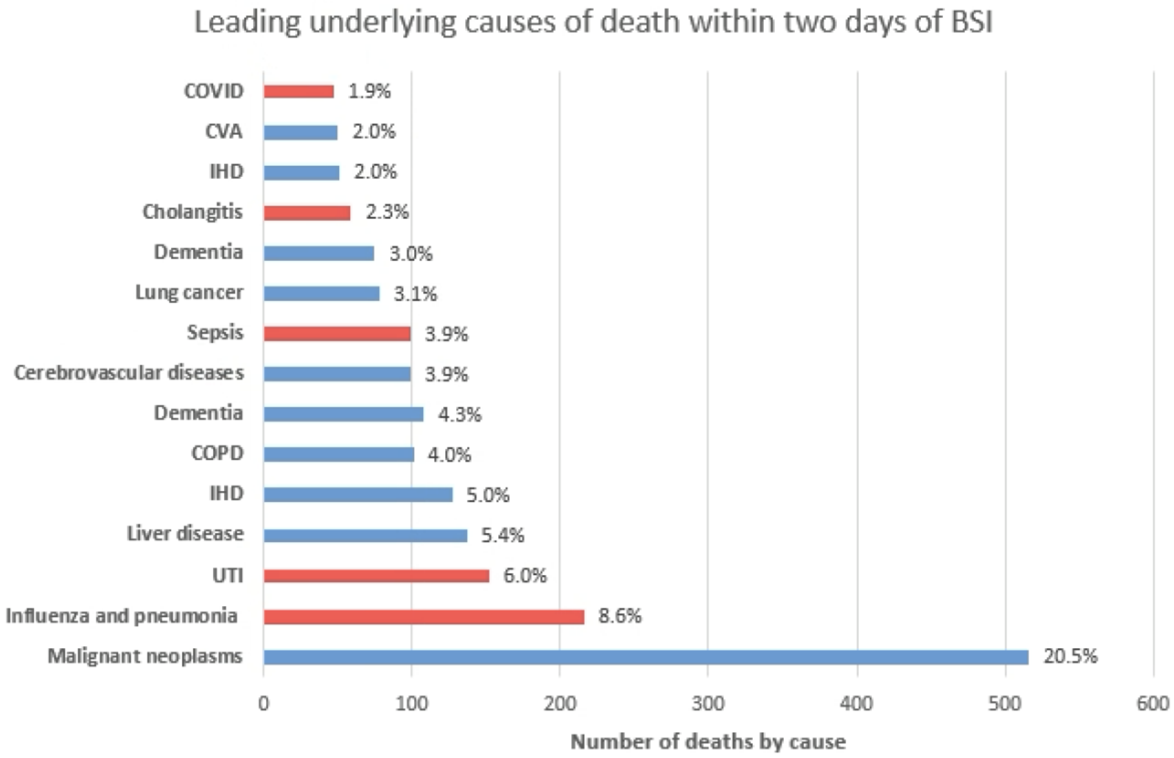

Leading underlying causes of death within two days of BSI
COVID
CVA
IHD
Cholangitis
Dementia
Lung cancer
Sepsis
Cerebrovascular diseases
Dementia
COPD
IHD
Liver disease
UTI
Influenza and pneumonia
Malignant neoplasms
1.9%
2.0%
2.0%
2.3%
3.0%
3.1%
3.9%
3.9%
4.3%
4.0%
5.0%
5.4%
6.0%
8.6%
20.5%
0
100
200
300
400
500
600
Number of deaths by cause
